# ASCVD risk refinement with NT-proBNP for statin allocation among low- and intermediate risk individuals

**DOI:** 10.1101/2024.04.09.24305587

**Authors:** Jelena Pavlović, Maryam Kavousi, M. Kamran Ikram, Daniel Bos, M. Arfan Ikram, Maarten J.G. Leening

## Abstract

**Background:** Statin trials targeting low- to intermediate risk individuals, namely MEGA, JUPITER, and HOPE-3, have demonstrated benefit of statin use for primary prevention of atherosclerotic cardiovascular disease (ASCVD), but are poorly reflected in guideline recommendations for primary prevention of ASCVD. N-terminal pro-B-type natriuretic peptide (NT-proBNP) may refine ASCVD risk in low-to intermediate risk individuals eligible for HOPE-3, JUPITER and MEGA, and aid statin initiation in low- to intermediate risk populations.

**Methods:** 5434 participants, aged 45 years and above from the prospective population-based Rotterdam Study, free of ASCVD, heart failure, and diabetes, were included between 1997 and 2008. Eligibility criteria for MEGA, JUPITER, and HOPE-3 trials were checked for each participant. ASCVD event rates, hazard ratios (HR), 5-year numbers needed to treat (NNT_5y_), and screen (NNS_5y_) per trial eligible population and NT-proBNP category (≤50, 50-100, and >100 pg/mL) were calculated.

**Results:** Median age was 61.6 years, 58.9% were women, median NT-proBNP was 60 pg/mL. The proportions of participants eligible for MEGA, JUPITER and HOPE-3 were 34.9%, 10.4% and 23.7%. Incidence rates per 1000 person-years for ASCVD were 10.4 (95%CI: 60.1-67.9) for MEGA, 16.8 (95%CI: 13.6-20.6) for JUPITER, and 12.1 (95%CI: 10.3-14) for HOPE-3. Adjusted HR in trial eligible individuals for NT-proBNP >100 pg/mL compared to ≤50 pg/mL level were 1.73 (95%CI: 1.21-2.47), 1.46 (95%CI: 0.80-2.66) and 1.50 (95%CI: 0.99-2.26), respectively. Estimated NNT_5y_ among trial eligible individuals with NT-proBNP levels >100 pg/mL based on high-intensity statin treatment, varied from 23 to 34 to prevent one ASCVD event, while NNS_5y_ ranged between 56 and 134.

**Conclusions:** NT-proBNP level >100 pg/mL identifies individuals at the highest ASCVD risk among low- to intermediate risk populations who are likely to benefit from statin treatment at acceptable NNT_5y_ and NNS_5y_.

**Clinical Perspective:**

1. What is New?

- NT-proBNP level >100 pg/mL can identify individuals at the highest ASCVD risk among low- to intermediate risk populations who are likely to benefit from statin treatment at acceptable numbers needed to treat and screen.
- Among apparently healthy individuals without prior ASCVD and diabetes mellitus, 60% was deemed to be at low- to intermediate risk by qualifying for at least one of three RCT based on the trial eligibility criteria, and one out four individuals had NT-proBNP level >100 pg/mL.
2. What are the Clinical Implications?

- NT-proBNP level can be used for risk refining in low- to intermediate risk individuals who are most likely to benefit from statin initiation for ASCVD primary prevention.

## INTRODUCTION

Management of blood cholesterol is one of the cornerstones of primary prevention of atherosclerotic cardiovascular disease (ASCVD).^1,2^ A variety of randomized clinical trials (RCT) have demonstrated the efficacy of statin treatment among individuals free of ASCVD.^3,4^ Three major RCT, namely the Management of Elevated Cholesterol in the Primary Prevention Group of Adult Japanese (MEGA)^5^, Justification for the Use of Statins in Prevention: Intervention Trial Evaluating Rosuvastatin (JUPITER)^6^, and Heart Outcomes Prevention Evaluation-3 (HOPE-3)^7,8^, have established the benefit of statin treatment in low- to intermediate risk individuals free of ASCVD.

We previously demonstrated that the findings of these three trials were poorly reflected in the US and European cholesterol treatment guideline recommendations.^9^ In addition, extending statin treatment recommendations to healthy individuals at low ASCVD risk raised concerns of statin overprescription.^10–12^ Hence, the latest US and European guidelines suggest using additional non-traditional risk factors, referred to as either risk enhancers or risk modifiers, to refine, and thus improve risk assessment in individuals at low- or intermediate ASCVD risk.^1,2^

In this regard, N-terminal pro-B-type natriuretic peptide (NT-proBNP) is a promising biomarker with potential to improve ASCVD risk assessment in low- to intermediate risk populations. NT-proBNP has been consistently associated with ASCVD risk,^13,14^ and is widely available and affordable,^15^ which makes it an appealing tool for ASCVD risk refinement in clinical practice. The value of NT-proBNP as a biomarker for risk refinement and statin allocation in primary prevention of ASCVD requires further evaluation.

In this study, we evaluate the application of NT-proBNP as a risk refining and decision-making tool in community-dwelling individuals meeting the eligibility criteria for three low- to intermediate risk statin RCT for primary prevention of ASCVD.

## METHODS

### Setting and Study Population

We used data from the Rotterdam Study, a prospective population-based cohort study drawn from the unselected population of a suburb in Rotterdam, designed to investigate etiology, preclinical phase, prognosis, and potential intervention targets for chronic diseases in mid- and late-life. The Rotterdam Study rationale and design have been described in detail previously.^16^

The Rotterdam Study recruited individuals in four subcohorts. Extensive examinations at baseline and follow-up visits were carried out every three to four years. NT-proBNP was measured in three subcohorts from 1997 to 2008. In total, 10,522 participants underwent examinations in these three visits combined, of which 9,326 participants had valid NT-proBNP measurement, informed consent, and follow-up (Figure S1).

All participants provided written informed consent to participate in the study and to have their information obtained from treating physicians. The Rotterdam Study has been approved by the Medical Ethics Committee of the Erasmus MC (registration number MEC 02.1015) and by the Dutch Ministry of Health, Welfare and Sport (Population Screening Act WBO, license number 1071272-159521-PG). The Rotterdam Study has been entered into the Netherlands National Trial Register (NTR; www.trialregister.nl) and into the WHO International Clinical Trials Registry Platform (ICTRP; www.who.int/ictrp/network/primary/en/) under shared catalogue number NTR6831.

### NT-proBNP Measurement

NT-proBNP was measured from blood samples stored at -80°C, collected in glass tubes with clot activator and serum separation gel. NT-proBNP was measured using an electrochemiluminescence immunoassay (Elecsys proBNP; Hoffman-La Roche Ltd., Basel, Switzerland) and Elecsys 2010 analyzer, as described previously.^17,18^

### Assessment of ASCVD

The main outcome was incident ASCVD, composed of myocardial infarction, coronary revascularization, non-hemorrhagic stroke, and ASCVD mortality.^19,20^ Events were adjudicated until January 1^st^, 2015. Follow-up and adjudication of outcome events was based on the general practitioners’ and medical specialists’ reports and verified by the study physicians, in detail described previously.^19,20^

### Assessment of Covariables

Information on medication use, smoking behavior, postmenopausal status, and medical history was collected by trained interviewers using a structured home interview. Anthropometric measures and laboratory testing were obtained during a visit to the research center.^13^ A detailed description of ASCVD risk factors assessment and used definitions for blood pressure, triglycerides, total, high-density and low-density cholesterol, glucose, c-reactive protein, thyroid-stimulating hormone, serum creatinine, estimated glomerular filtration rate, alanine aminotransferase, presence of diabetes mellitus and family history of premature coronary heart disease are provided in the Supplemental Methods.

### Statin Trial Eligibility

We followed eligibility criteria for three double blind placebo-controlled RCT on statin use in primary prevention targeted at low- to intermediate risk individuals with hard ASCVD end points: MEGA,^5,21^ JUPITER,^6^ and HOPE-3^7^ (Table 1, Table S1).

**Table 1.**
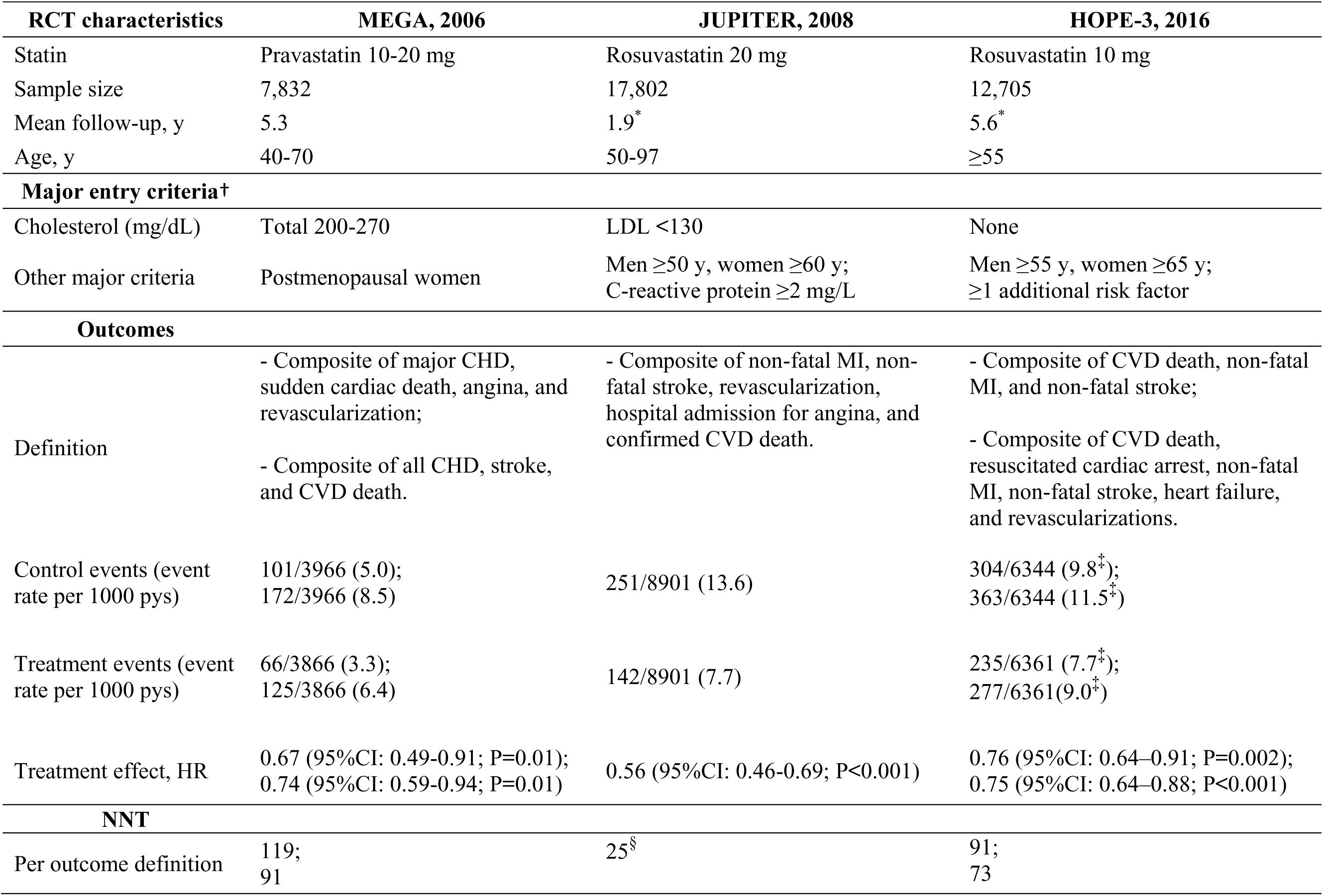
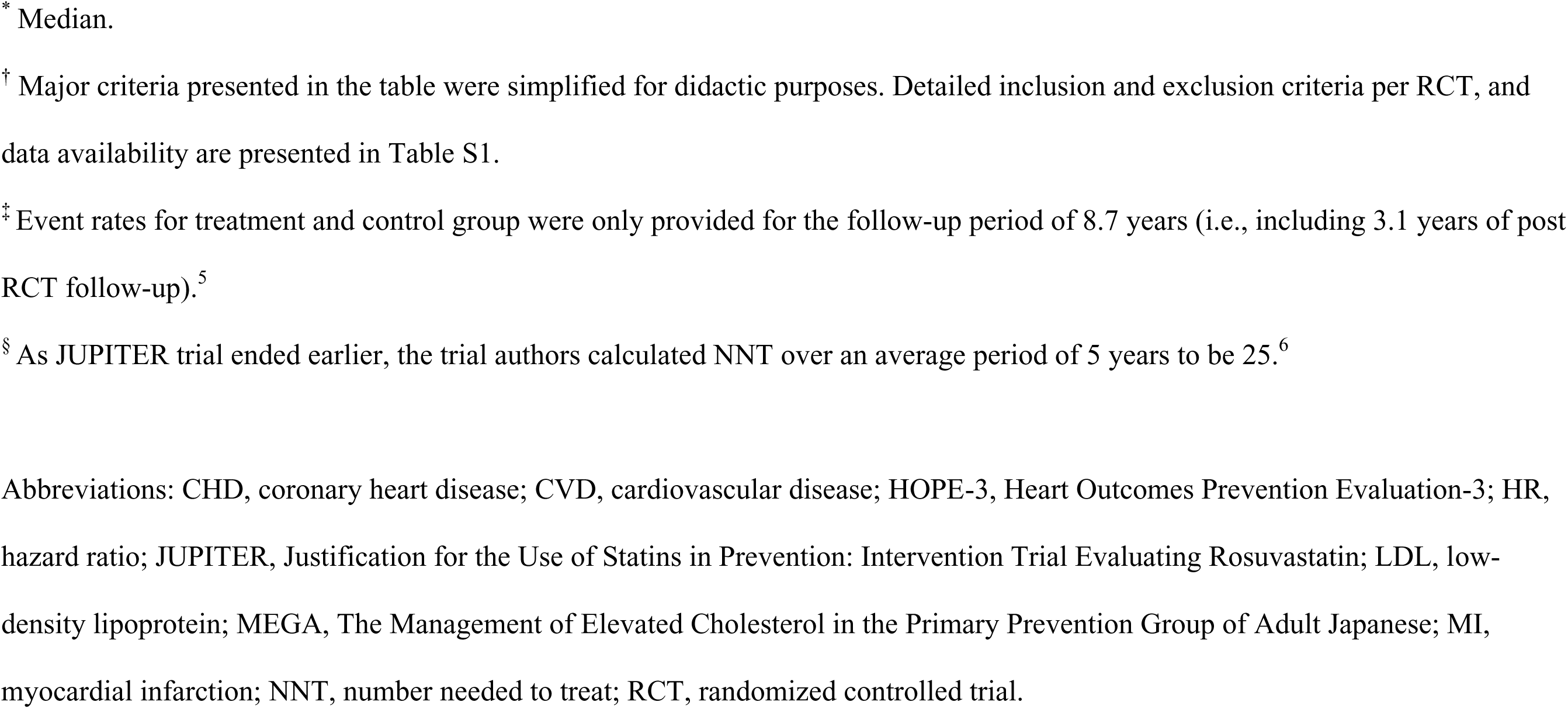
Overview of the clinical trial characteristics.

| **RCT characteristics** | **MEGA, 2006** | **JUPITER, 2008** | **HOPE-3, 2016** |
| --- | --- | --- | --- |
| Statin | Pravastatin 10-20 mg | Rosuvastatin 20 mg | Rosuvastatin 10 mg |
| Sample size | 7,832 | 17,802 | 12,705 |
| Mean follow-up, y | 5.3 | 1.9* | 5.6* |
| Age, y | 40-70 | 50-97 | ≥55 |
| **Major entry criteria<sup>†</sup>** |  |  |  |
| Cholesterol (mg/dL) | Total 200-270 | LDL <130 | None |
| Other major criteria | Postmenopausal women | Men ≥50 y, women ≥60 y;<br>C-reactive protein ≥2 mg/L | Men ≥55 y, women ≥65 y;<br>≥1 additional risk factor |
| **Outcomes** |  |  |  |
| Definition | - Composite of major CHD, sudden cardiac death, angina, and revascularization;<br>- Composite of all CHD, stroke, and CVD death. | - Composite of non-fatal MI, non-fatal stroke, revascularization, hospital admission for angina, and confirmed CVD death. | - Composite of CVD death, non-fatal MI, and non-fatal stroke;<br>- Composite of CVD death, resuscitated cardiac arrest, non-fatal MI, non-fatal stroke, heart failure, and revascularizations. |
| Control events (event rate per 1000 pys) | 101/3966 (5.0);<br>172/3966 (8.5) | 251/8901 (13.6) | 304/6344 (9.8<sup>‡</sup>);<br>363/6344 (11.5<sup>‡</sup>) |
| Treatment events (event rate per 1000 pys) | 66/3866 (3.3);<br>125/3866 (6.4) | 142/8901 (7.7) | 235/6361 (7.7<sup>‡</sup>);<br>277/6361 (9.0<sup>‡</sup>) |
| Treatment effect, HR | 0.67 (95%CI: 0.49-0.91; P=0.01);<br>0.74 (95%CI: 0.59-0.94; P=0.01) | 0.56 (95%CI: 0.46-0.69; P<0.001) | 0.76 (95%CI: 0.64–0.91; P=0.002);<br>0.75 (95%CI: 0.64–0.88; P<0.001) |
| **NNT** |  |  |  |
| Per outcome definition | 119;<br>91 | 25<sup>§</sup> | 91;<br>73 |

\* Median.
† Major criteria presented in the table were simplified for didactic purposes. Detailed inclusion and exclusion criteria per RCT, and data availability are presented in Table S1.
‡ Event rates for treatment and control group were only provided for the follow-up period of 8.7 years (i.e., including 3.1 years of post RCT follow-up).<sup>5</sup>
§ As JUPITER trial ended earlier, the trial authors calculated NNT over an average period of 5 years to be 25.<sup>6</sup>
Abbreviations: CHD, coronary heart disease; CVD, cardiovascular disease; HOPE-3, Heart Outcomes Prevention Evaluation-3; HR, hazard ratio; JUPITER, Justification for the Use of Statins in Prevention: Intervention Trial Evaluating Rosuvastatin; LDL, low- density lipoprotein; MEGA, The Management of Elevated Cholesterol in the Primary Prevention Group of Adult Japanese; MI, myocardial infarction; NNT, number needed to treat; RCT, randomized controlled trial.

Inclusion criteria among the RCT varied due to differences in design and hypothesis. Major RCT inclusion criteria included: age, sex, lipid levels, C-reactive protein levels, history of diabetes mellitus, blood pressure, and postmenopausal status for women (Table 1, Table S1). Data on a few minor exclusion criteria were not available in the Rotterdam Study, such as creatine phosphokinase levels, illicit drug abuse, and rare medical conditions (Table S1).

Finally, to select apparently healthy individuals and emulate eligibility criteria for three RCT (Figure S1), in the present analysis we included a total of 5434 participants over 45 years of age, without a history of diabetes mellitus, ASCVD, and heart failure, with low-density lipoprotein (LDL) cholesterol levels below 190 mg/dL (4.9 mmol/L), not using a statin, and without uncontrolled risk factors. The uncontrolled risk factors in essence reflected minor exclusion criteria defined by three RCT, such as uncontrolled hypertension, hypothyroidism, history of chronic liver disease, use of systemic corticosteroids or cyclosporin, and high levels of serum creatinine and triglycerides, as defined in the Supplemental Methods. History of ASCVD was defined as any of the following: myocardial infarction, coronary or other arterial revascularization procedure, stroke, transient ischemic attack, or repeated prescription of nitrates (as a proxy for angina pectoris).^19,20,22,23^ We excluded individuals with age-specific levels of NT-proBNP indicative of underlying heart failure (if age <50 years and NT-proBNP >450 pg/mL, or age 50-75 years and NT-proBNP >900 pg/mL, or age >75y and NT-proBNP >1800 pg/mL).^24^

### Statistical Analysis

First, we determined the cardiovascular risk factor profiles for each trial, and proportions of the study population meeting eligibility criteria for MEGA, JUPITER, and HOPE-3. In addition, we divided the trial eligible populations using three NT-proBNP thresholds (≤50 pg/mL, 50-100 pg/mL, and >100 pg/mL).^25^

Next, we calculated incidence rates for ASCVD events per 1000 person-years in the entire study population, trial eligible subgroups, and by NT-proBNP categories.

To evaluate the risk refining properties of the three NT-proBNP categories, we applied Cox regression models. Specifically, we analyzed associations of continuous natural-log-transformed NT-proBNP, and NT-proBNP categories in relation to the incidence of ASCVD events in the total population as well as in trial eligible subgroups. The reference for comparison was the lowest NT-proBNP category (≤50 pg/mL). We report adjusted hazard ratios (HR) for two models: model 1, adjusted for age and sex, and model 2, additionally adjusted for traditional risk factors: current smoking, systolic blood pressure, total and high-density lipoprotein cholesterol.

To explore the potential yield of NT-proBNP as a risk refining and statin allocation tool in the context of ASCVD primary prevention, we calculated 5-year numbers needed to treat (NNT_5y_) and numbers needed to screen (NNS_5y_) for each NT-proBNP category.

Based on the observed ASCVD event rates and the anticipated risk reduction with statin therapy, we calculated NNT_5y_. We calculated NNT_5y_ for 30% and 50% LDL cholesterol reduction, corresponding to treatment with moderate- and high-intensity statins, respectively.^26^ For primary prevention, expected relative risk reduction of 25% in major ASCVD events was used for each 38.7 mg/dL (1.0 mmol/L) of LDL cholesterol lowering.^3^ We calculated NNT_5y_ using the following formula: NNT_5y_ = 1 / (5 * [observed ASCVD incidence rate] * [observed median LDL in mmol/L] * [anticipated proportional LDL reduction] * [anticipated ASCVD relative risk reduction]).

NNS_5y_ to prevent one ASCVD event were calculated based on the NNT_5y_ and NT-proBNP level distributions for trial eligible subgroups. NNS_5y_ were calculated by multiplying NNT_5y_ with inverse probability of the prevalence of the NT-proBNP threshold, in total population and trial eligible subgroups.

Sensitivity analyses restricted to a composite coronary heart disease (CHD) outcome were performed and presented the corresponding results in the Supplemental Material. The CHD outcome was defined as a composite of myocardial infarction, coronary revascularization, and CHD mortality.^19^ For CHD, the relative risk reduction of 29% in CHD events was used for each 38.7 mg/dL (1.0 mmol/L) of LDL cholesterol lowering to calculate respective NNT_5y_ and NNS_5y_.^3^

We had complete data for 95.7% of traditional cardiovascular risk factors. Missing values were handled by single imputation using an expectation-maximization algorithm, separately for each sex, using age and all other traditional risk factors.^27^ We used IBM SPSS Statistics version 25.0 (IBM Corp, Armonk, NY, USA) and R version 4.0.4 (R Foundation for Statistical Computing, Vienna, Austria) for all analyses.

## RESULTS

Median age in the total population was 61.6 years and 58.9% were women. The median NT-proBNP was 60 pg/mL (Table 2). MEGA eligible individuals were younger and had lower median NT-proBNP levels compared to JUPITER and HOPE-3 eligible individuals. The HOPE-3 eligible population had the most unfavorable lipid profile.

**Table 2.**
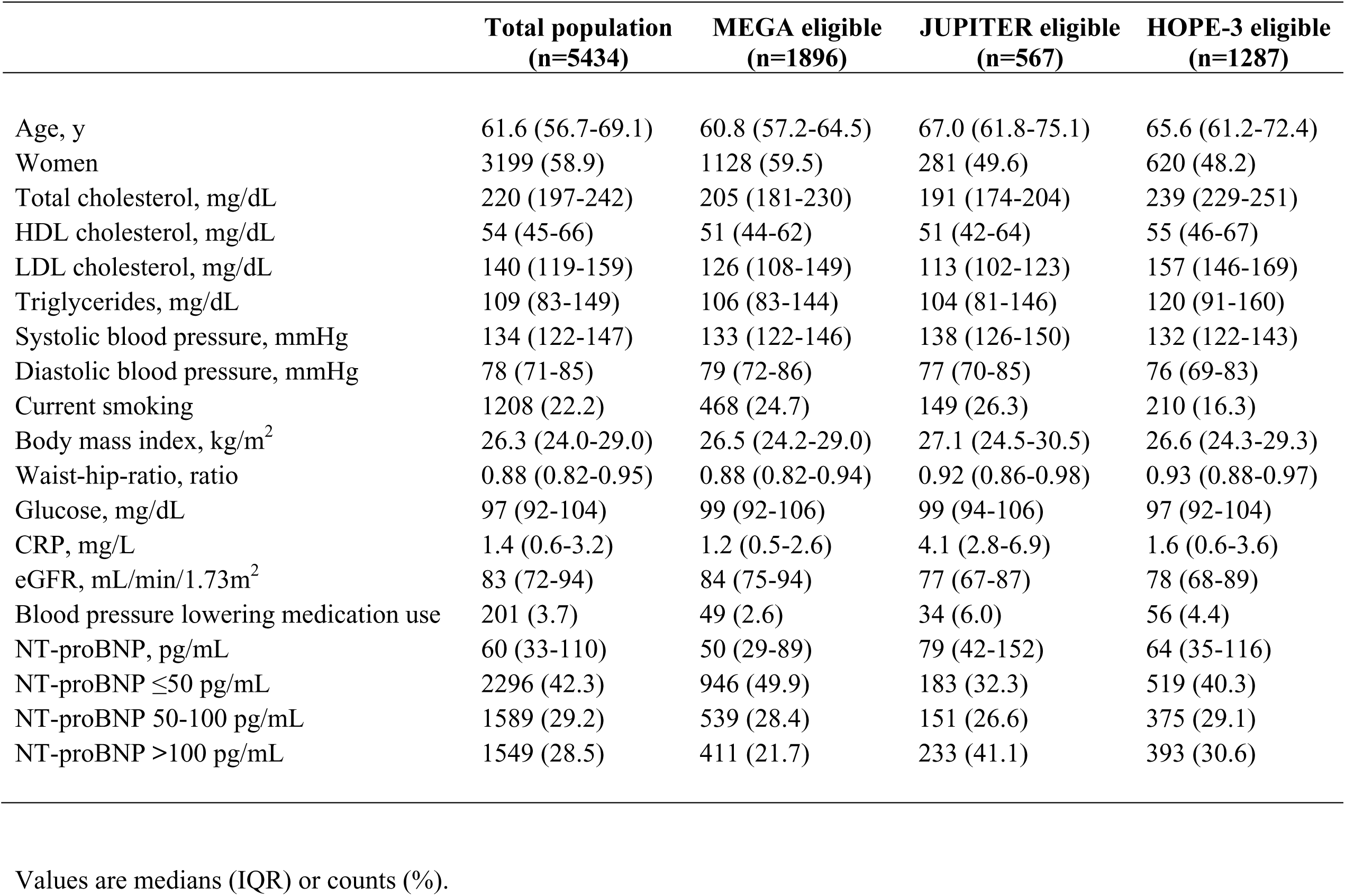
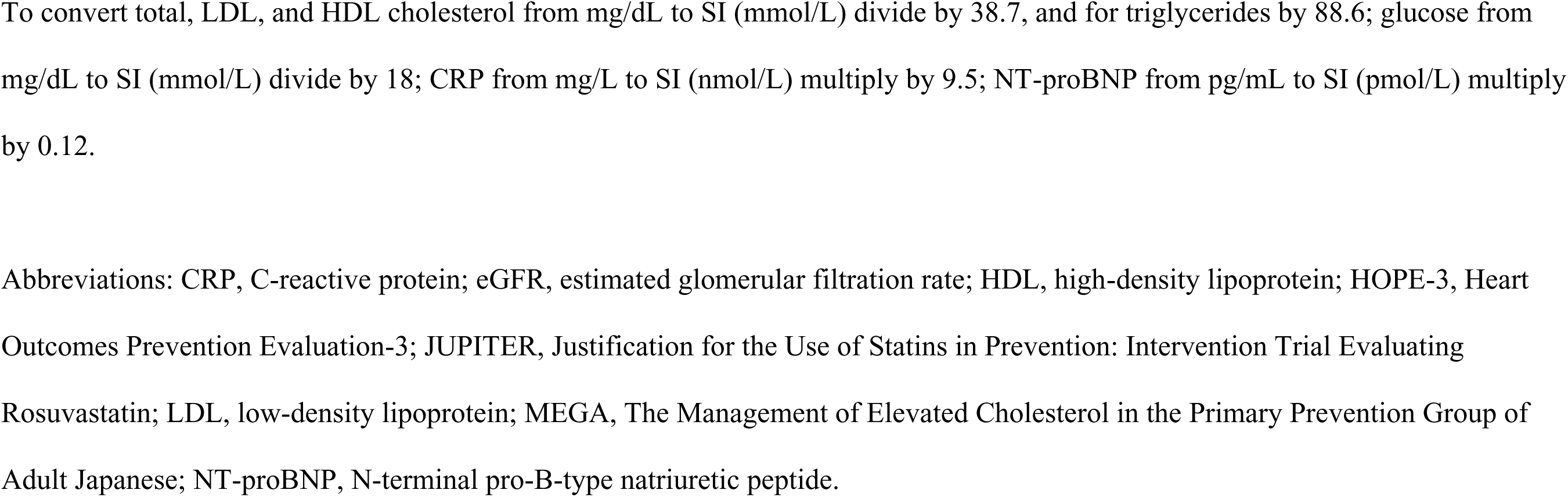
Population characteristics.

|  | **Total population**<br>**(n=5434)** | **MEGA eligible**<br>**(n=1896)** | **JUPITER eligible**<br>**(n=567)** | **HOPE-3 eligible**<br>**(n=1287)** |
| --- | --- | --- | --- | --- |
| Age, y | 61.6 (56.7-69.1) | 60.8 (57.2-64.5) | 67.0 (61.8-75.1) | 65.6 (61.2-72.4) |
| Women | 3199 (58.9) | 1128 (59.5) | 281 (49.6) | 620 (48.2) |
| Total cholesterol, mg/dL | 220 (197-242) | 205 (181-230) | 191 (174-204) | 239 (229-251) |
| HDL cholesterol, mg/dL | 54 (45-66) | 51 (44-62) | 51 (42-64) | 55 (46-67) |
| LDL cholesterol, mg/dL | 140 (119-159) | 126 (108-149) | 113 (102-123) | 157 (146-169) |
| Triglycerides, mg/dL | 109 (83-149) | 106 (83-144) | 104 (81-146) | 120 (91-160) |
| Systolic blood pressure, mmHg | 134 (122-147) | 133 (122-146) | 138 (126-150) | 132 (122-143) |
| Diastolic blood pressure, mmHg | 78 (71-85) | 79 (72-86) | 77 (70-85) | 76 (69-83) |
| Current smoking | 1208 (22.2) | 468 (24.7) | 149 (26.3) | 210 (16.3) |
| Body mass index, kg/m<sup>2</sup> | 26.3 (24.0-29.0) | 26.5 (24.2-29.0) | 27.1 (24.5-30.5) | 26.6 (24.3-29.3) |
| Waist-hip-ratio, ratio | 0.88 (0.82-0.95) | 0.88 (0.82-0.94) | 0.92 (0.86-0.98) | 0.93 (0.88-0.97) |
| Glucose, mg/dL | 97 (92-104) | 99 (92-106) | 99 (94-106) | 97 (92-104) |
| CRP, mg/L | 1.4 (0.6-3.2) | 1.2 (0.5-2.6) | 4.1 (2.8-6.9) | 1.6 (0.6-3.6) |
| eGFR, mL/min/1.73m<sup>2</sup> | 83 (72-94) | 84 (75-94) | 77 (67-87) | 78 (68-89) |
| Blood pressure lowering medication use | 201 (3.7) | 49 (2.6) | 34 (6.0) | 56 (4.4) |
| NT-proBNP, pg/mL | 60 (33-110) | 50 (29-89) | 79 (42-152) | 64 (35-116) |
| NT-proBNP ≤50 pg/mL | 2296 (42.3) | 946 (49.9) | 183 (32.3) | 519 (40.3) |
| NT-proBNP 50-100 pg/mL | 1589 (29.2) | 539 (28.4) | 151 (26.6) | 375 (29.1) |
| NT-proBNP >100 pg/mL | 1549 (28.5) | 411 (21.7) | 233 (41.1) | 393 (30.6) |

Values are medians (IQR) or counts (%).
To convert total, LDL, and HDL cholesterol from mg/dL to SI (mmol/L) divide by 38.7, and for triglycerides by 88.6; glucose from mg/dL to SI (mmol/L) divide by 18; CRP from mg/L to SI (nmol/L) multiply by 9.5; NT-proBNP from pg/mL to SI (pmol/L) multiply by 0.12.
Abbreviations: CRP, C-reactive protein; eGFR, estimated glomerular filtration rate; HDL, high-density lipoprotein; HOPE-3, Heart Outcomes Prevention Evaluation-3; JUPITER, Justification for the Use of Statins in Prevention: Intervention Trial Evaluating Rosuvastatin; LDL, low-density lipoprotein; MEGA, The Management of Elevated Cholesterol in the Primary Prevention Group of Adult Japanese; NT-proBNP, N-terminal pro-B-type natriuretic peptide.

Overall, 1896 (34.9%, 95%CI: 33.6-36.2), 567 (10.4%, 95%CI: 9.6-11.3), and 1287 (23.7%, 95%CI: 22.6-24.8) individuals of the total study population fitted the eligibility criteria for MEGA, JUPITER, and HOPE-3, respectively (Figure 1). JUPITER had the highest proportion of eligible individuals with NT-proBNP levels >100 pg/mL (41.1%), as compared to MEGA (21.7%) and HOPE-3 (30.6%) eligible individuals (Table 2).

**Figure 1.**
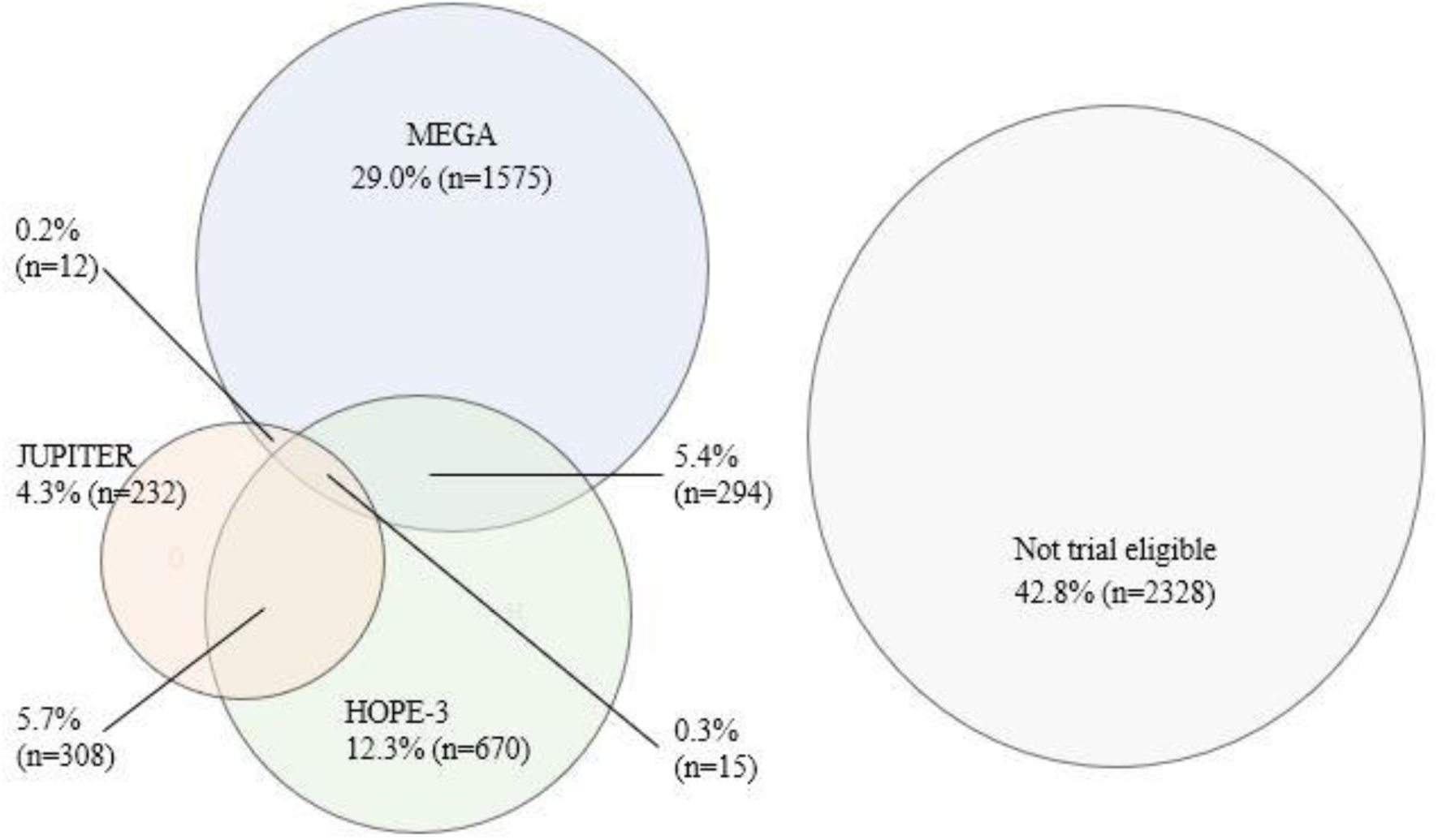
General population eligibility for statin trials in primary prevention of ASCVD targeting low- and intermediate risk individuals. Abbreviations: ASCVD, atherosclerotic cardiovascular disease; HOPE-3, Heart Outcomes Prevention Evaluation-3; JUPITER, Justification for the Use of Statins in Prevention: Intervention Trial Evaluating Rosuvastatin; MEGA, The Management of Elevated Cholesterol in the Primary Prevention Group of Adult Japanese.

In total population, during a mean (SD) follow-up of 9.9 (4.2) years, 681 first ASCVD events in total were observed, corresponding to an ASCVD event rate of 12.6 (95%CI: 11.7-13.6) per 1000 person-years. Among trial eligible individuals with NT-proBNP levels >100 pg/mL, JUPITER had the highest ASCVD incidence rate (24.0 per 1000 person-years), and MEGA the lowest (15.0 per 1000 person-years) (Table S2).

NT-proBNP >100 pg/mL was significantly associated with ASCVD incidence after adjusting for traditional risk factors (model 2) in the total study population and among individuals eligible for MEGA (Figures 2). Multivariable adjusted HR for ASCVD events comparing NT-proBNP levels of >100 pg/mL and ≤50 pg/mL were 1.56 (95%CI: 1.27-1.91; P<0.001) in the total population, 1.72 (95%CI: 1.20-2.45; P=0.003) among MEGA eligible, 1.46 (95%CI: 0.80-2.67; P=0.213) among JUPITER eligible, and 1.50 (95%CI: 1.00-2.27; P=0.052) among HOPE-3 eligible individuals (Figure 2). Adjustment for traditional risk factors only marginally affected the HR (Table S3). The ASCVD free survival was lowest across all trial eligible individuals when NT-proBNP levels were >100 pg/mL (Figure 3).

**Figure 2.**
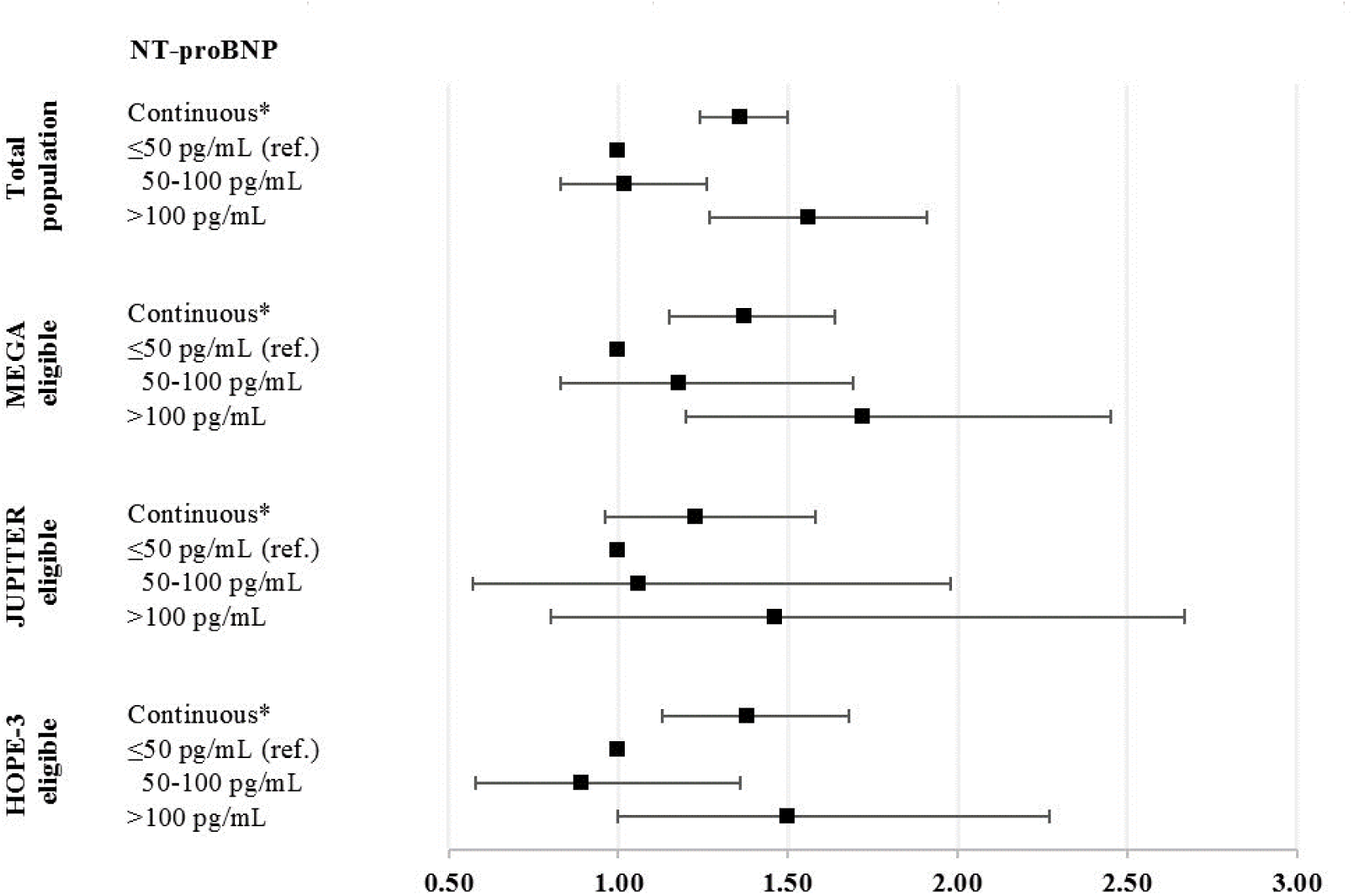
Adjusted hazard ratios for incident ASCVD. * Natural-log-transformed NT-proBNP values. Adjusted hazard ratios (model 2) for ASCVD with 95% confidence intervals per natural-log-transformed NT-proBNP values, comparing two NT-proBNP categories with NT-proBNP ≤50 pg/mL (reference group) in total population and per trial eligible subpopulations. Abbreviations: ASCVD, atherosclerotic cardiovascular disease; HDL, high-density lipoprotein; HOPE-3, Heart Outcomes Prevention Evaluation-3; JUPITER, Justification for the Use of Statins in Prevention: Intervention Trial Evaluating Rosuvastatin; MEGA, The Management of Elevated Cholesterol in the Primary Prevention Group of Adult Japanese; NT-proBNP, N-terminal pro-B-type natriuretic peptide.

**Figure 3.**
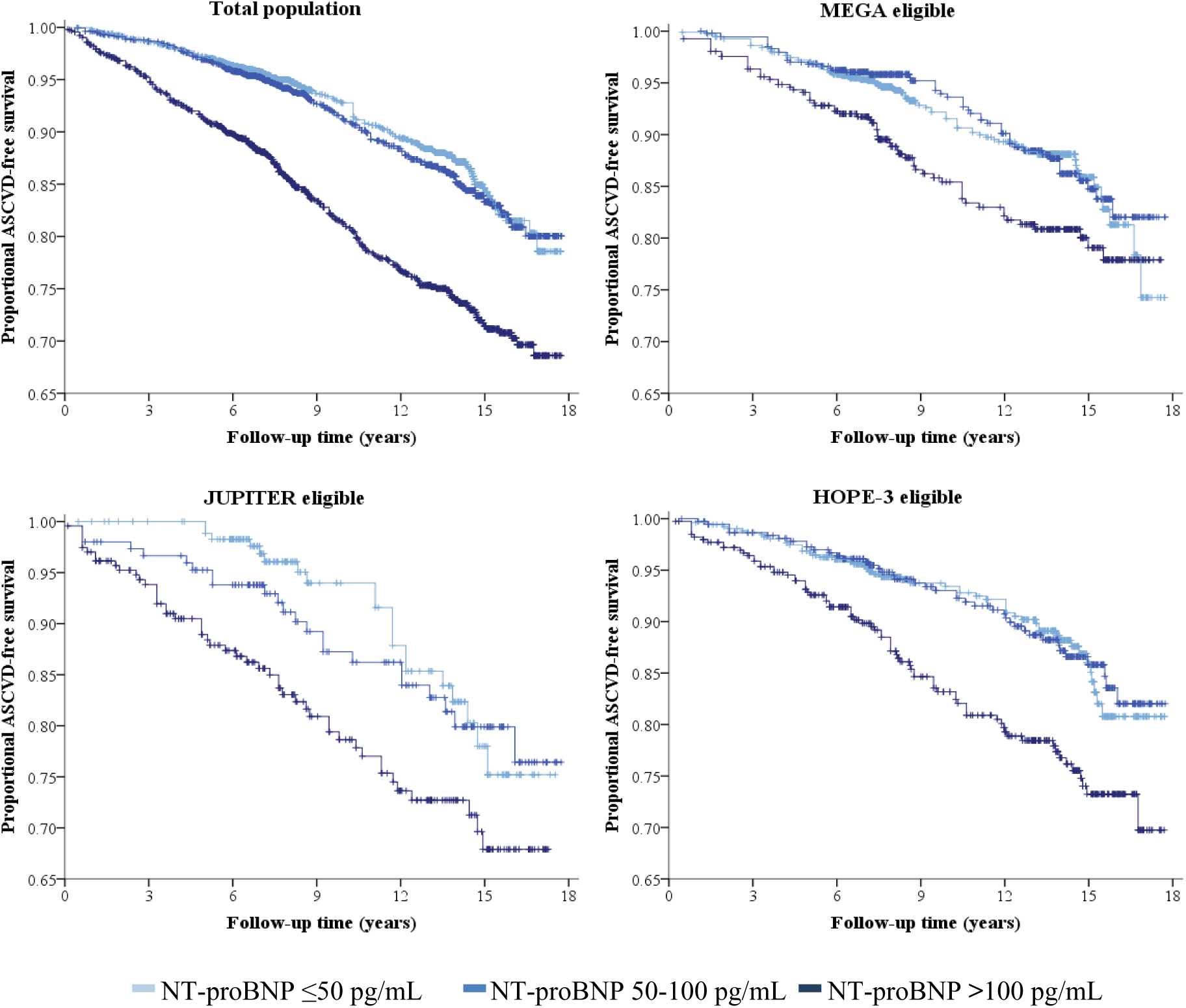
ASCVD free survival curves by NT-proBNP level and trial eligibility. Abbreviations: ASCVD, atherosclerotic cardiovascular disease; HOPE-3, Heart Outcomes Prevention Evaluation-3; JUPITER, Justification for the Use of Statins in Prevention: Intervention Trial Evaluating Rosuvastatin; MEGA, The Management of Elevated Cholesterol in the Primary Prevention Group of Adult Japanese; NT-proBNP, N-terminal pro-B-type natriuretic peptide.

Estimated NNT_5y_ to avoid one ASCVD event were lowest for JUPITER eligible individuals, except for those with NT-proBNP levels ≤50 pg/mL (Figure 4, Table S4). Among MEGA, JUPITER and HOPE-3 eligible with NT-proBNP levels >100 pg/mL, 48, 39 and 56 individuals would need to be treated with moderate-intensity statins for 5 years to prevent one ASCVD event. With high-intensity statins, NNT_5y_ were 29 for MEGA, 23 for JUPITER, and 34 for HOPE-3.

**Figure 4.**
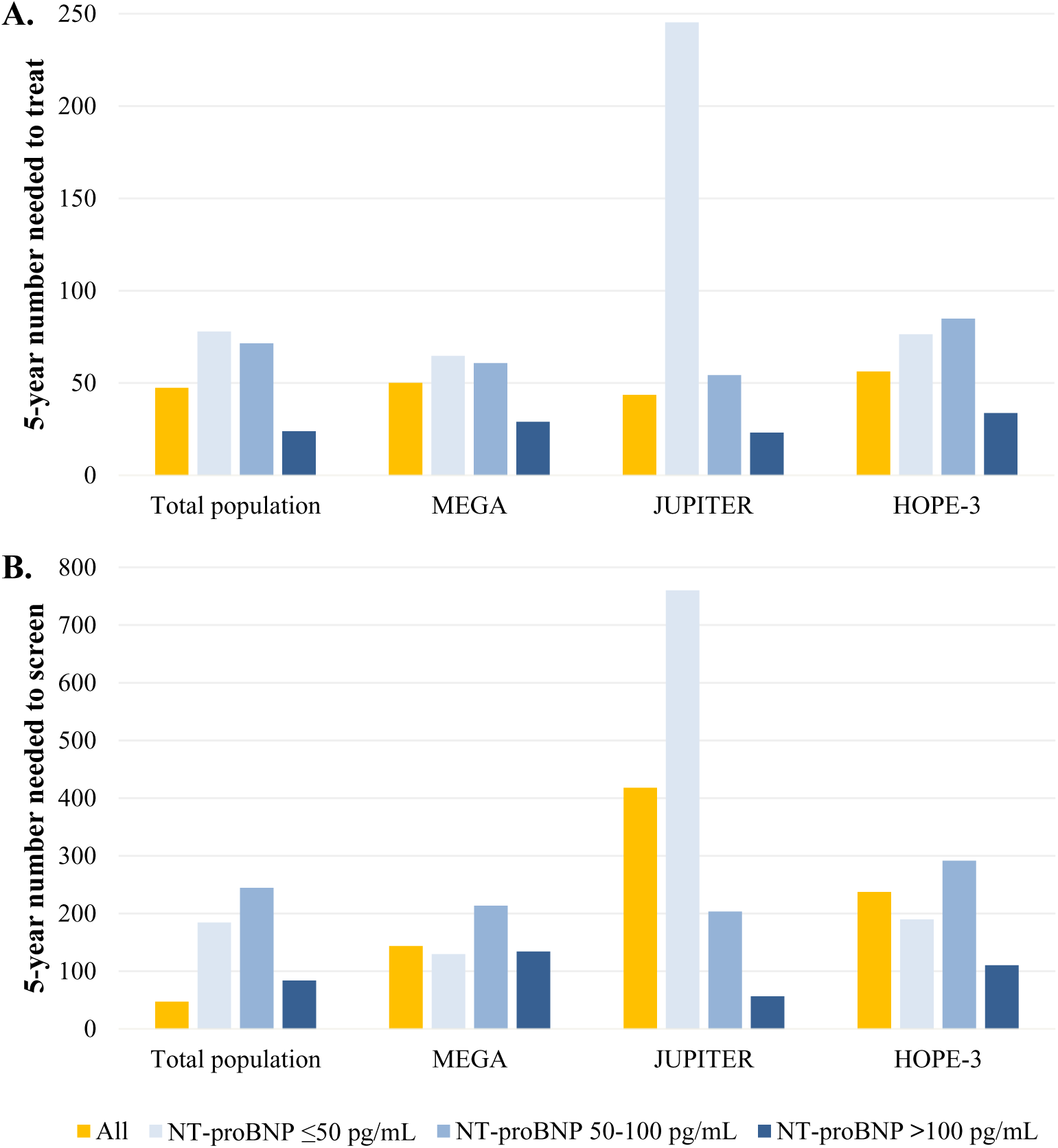
Numbers needed to treat and screen for five years to avoid a single ASCVD event with high-intensity statins. Panel A, Numbers needed to treat for five years to avoid a single ASCVD event with high-intensity statins, by NT-proBNP level. Panel B, Numbers needed to screen for five years to avoid a single ASCVD event with high-intensity statins, by NT-proBNP level. Abbreviations: ASCVD, atherosclerotic cardiovascular disease; HOPE-3, Heart Outcomes Prevention Evaluation-3; JUPITER, Justification for the Use of Statins in Prevention: Intervention Trial Evaluating Rosuvastatin; MEGA, The Management of Elevated Cholesterol in the Primary Prevention Group of Adult Japanese; NT-proBNP, N-terminal pro-B-type natriuretic peptide.

Among individuals with NT-proBNP levels >100 pg/mL for MEGA, JUPITER, and HOPE-3 eligible, NNS_5y_ based on the NNT_5y_ with high-intensity statins were 134, 56 and 111, respectively (Figure 4). Estimated NNT_5y_ and NNS_5y_ were lower for high-intensity statin treatment compared to moderate-intensity statin treatment (Table S4).

Results for CHD outcomes followed a similar pattern to those for ASCVD (Tables S5 and S6, Figures S2 and S3). Estimated NNT_5y_ and NNS_5y_ to prevent an ASCVD event were lower than those needed to prevent a CHD event (Table S4).

## DISCUSSION

NT-proBNP levels >100 pg/mL identify individuals at an increased risk of ASCVD among individuals at low- to intermediate risk who are eligible for statin RCT for primary prevention of ASCVD.^5–7^ We found comparable numbers needed to treat to those reported by the three original trials. Our results imply that NT-proBNP could be considered as a risk refining tool among individuals at low- to intermediate risk for primary prevention of ASCVD.

The percentages of eligibility for three statin primary prevention trials were 34.9% for MEGA, 10.4% for JUPITER and 23.7% for HOPE-3, with minimal overlap between trials, meaning that these trials differentially selected low- to intermediate risk profiles in the general population free of ASCVD. Overall, 57.2% of the general population sample free of ASCVD and diabetes, was eligible for at least one of these three RCT, meaning that for these individuals, the evidence of the statin efficacy to prevent ASCVD is established. This finding emphasizes the broad evidence base for statin use in primary prevention of ASCVD, which is in stark contrast with the small number of selected very high-risk individuals that is currently strongly recommended statin therapy in the most recent European prevention guidelines.^28^ Although NT-proBNP >100 pg/mL was significantly associated with ASCVD incidence in the total study population and MEGA eligible individuals (Figures 2), but not among JUPITER and HOPE-3 eligible, the association was comparable across trial eligible populations. The lack of significant association might be explained with limited number of events during follow-up, given that substantially fewer individuals qualified for JUPITER and HOPE-3 compared to MEGA (Table S3).

In this study we used NT-proBNP as a tool to refine ASCVD risk to identify low- to intermediate risk individuals for whom evidence supports preventive statin treatment. To examine the yield of statin treatment in the trial eligible population we calculated NNT_5y_ and NNS_5y_. NNT is commonly used in clinical trials to assess the acceptability of the investigated intervention.^29,30^ Generally, the lower the NNT, the more acceptable the intervention is. NNT over approximately five years of statin treatment reported by the three evaluated statin trials ranged from 25 to 119 (Table 1).^5–8^ In the present study, we report comparable NNT_5y_ varying from 23 to 34 for ASCVD for high-intensity statins to prevent one event among trial eligible individuals with NT-proBNP levels >100 pg/mL. In addition, NNS provides an estimate of the number of individuals that need to undergo NT-proBNP testing to identify individuals at NT-proBNP levels >100 pg/mL, and subsequently prevent one ASCVD event in the coming five years. In our study, among trial eligible individuals, NNS_5y_ ranged between 56 to 134 with high-intensity statins. Our results indicate that NT-proBNP level >100 pg/mL can identify individuals at the highest ASCVD risk among low- to intermediate risk populations who are likely to benefit from statin treatment at acceptable NNT_5y_ and NNS_5y_. Ultimately, whether these estimated NNT_5y_ and NNS_5y_ are acceptable for clinical practice is highly dependent on the healthcare system. Although NNT has its limitations,^30^ it can inform a shared decision-making between patient and healthcare provider regarding the pros and cons of statin use in primary prevention of ASCVD.

## Implications

As apparently healthy individuals at low- to intermediate ASCVD risk are more likely to have ASCVD risk around the decision thresholds, in this population NT-proBNP can further inform decision regarding the allocation of statin treatment. We demonstrated that statin initiation in this population based on NT-proBNP level of >100 pg/mL could be acceptable in terms of how many individuals would need to be screened and treated with statins to avoid one ASCVD event. Current US and European ASCVD prevention guidelines do not yet recommend the use of NT-proBNP as a risk enhancer or modifier for risk stratification.^1,2^ In contrast, NT-proBNP has been endorsed in the latest iteration of the ESC guidelines for peri-operative cardiac care to identify apparently healthy individuals at increased risk for peri-operative cardiac events.^31^ Furthermore, compared to imaging methods endorsed by the US and European ASCVD prevention guidelines, the price of a single NT-proBNP measurement is considerably lower. In addition, NT-proBNP testing could be easily integrated into routine clinical practice. Our results suggest that the application of NT-proBNP as a risk refining tool for primary prevention warrants further evaluation, including defining optimal age- and sex-specific thresholds for clinical decision-making in ASCVD risk refinement. For example, NT-proBNP levels are higher in women,^32^ NT-proBNP increases with age,^24^ and co-morbidities further affect levels of NT-proBNP.^33^

### Limitations

Specific limitations of our study need to be addressed. As most of the Rotterdam Study population is of European descent (95.7% white), our findings require validation in other ethnicities. Although large differences across ethnicities are not expected, it is relevant to establish generalizability in different populations. Due to the healthy volunteer effect observed in the Rotterdam Study cohort, the prevalence of cardiovascular risk factors and ASCVD incidence rates during follow-up are likely somewhat lower than in real-world data setting.^34^ Therefore, our results need to be interpreted with caution when extrapolating findings to the broader population accessing healthcare services.

### Conclusions

NT-proBNP levels >100 pg/mL identify individuals at the highest ASCVD risk among low- to intermediate risk populations who are likely to benefit from statin treatment at acceptable NNT_5y_ and NNS_5y_. Almost 60% of the general population free of ASCVD and diabetes was eligible for at least one of three statin RCT, and one out four had NT-proBNP levels >100 pg/mL.

## Data Availability

The data supporting the findings of this study are available from the corresponding author upon reasonable request.

## Acknowledgments

The dedication, commitment, and contribution of inhabitants, general practitioners, and pharmacists to the Rotterdam Study are gratefully acknowledged.

## Supplemental Material

Supplemental Methods

Tables S1-S6

Figures S1-S3

References 35-36

## Non-standard Abbreviations and Acronyms

ASCVD: atherosclerotic cardiovascular disease
CHD: coronary heart disease
HOPE-3: Heart Outcomes Prevention Evaluation-3 Trial
JUPITER: Justification for the Use of Statins in Prevention: Intervention Trial Evaluating Rosuvastatin
MEGA: The Management of Elevated Cholesterol in the Primary Prevention Group of Adult Japanese Trial
NNS: number needed to screen
NNT: number needed to treat
NT-proBNP: N-terminal pro-B-type natriuretic peptide
RCT: randomized controlled trial

